# A Histological Assessment Tool for Breast Implant Capsules Validated in 480 Patients with and without Capsular Contracture

**DOI:** 10.1101/2023.04.23.23289002

**Authors:** Andreas Larsen, Adam M Timmermann, Mikela Kring, Tim K Weltz, Mathias Ørholt, Peter Vester-Glowinski, Jens Jørgen Elberg, Jesper Trillingsgaard, Louise V Mielke, Lisbet R Hölmich, Tine E Damsgaard, Anne Roslind, Mikkel Herly

## Abstract

**Background:** Capsular contracture is a severe complication to breast implants, but the pathophysiology remains unclear, and consensus is lacking on how to analyze implant capsules histologically. In this study, we developed and validated a histological semiquantitative assessment tool for analyzing breast implant capsules.

**Methods:** Biopsies of breast implant capsules from 480 patients who had undergone breast augmentation or reconstruction were collected and stained with hematoxylin and eosin. First, biopsies from 100 patients were used to select parameters for the semiquantitative assessment tool. Then, biopsies from the remaining 380 patients were used to determine intra- and interobserver agreement of two blinded observers and agreement with a pathologist. Finally, we tested the association between the parameters and capsular contracture.

**Results:** The histological assessment tool consisted of ten parameters assessed on a two-, three-, or four-point scale. Intra- and interobserver agreement was almost perfect (0.83 and 0.80), and agreement with the pathologist was substantial (0.67). Inflammatory infiltration (*p*<0.01), thickness of the collagen layer (*p*<0.0001), fiber organization (*p*<0.01) and calcification (*p*<0.001) were significantly correlated with capsular contracture.

**Conclusions:** This is the first validated histological assessment tool for breast implant capsules that provides an objective and precise assessment of hematoxylin and eosin-stained sections of breast implant capsule biopsies which can be used in research and clinical practice.

**Level of evidence:** No Level Assigned

## Introduction

Capsular contracture is one of the most common severe complications of breast implants. Within 10 years after surgery, capsular contracture affects 3.6-19%[1–5] of women after breast augmentation and 3.3-25%[1, 3–6] of women after implant-based breast reconstruction. The severity of capsular contracture is most often clinically evaluated using the Baker Classification System, which consists of a four-point scale based on the firmness and distortion of the breast.[7, 8] Despite the importance of capsular contracture, its pathogenesis is largely unknown.[9, 10] Previous studies suggest that capsular contracture is caused by a low-grade inflammatory response in the tissue around the breast implant, which results in excessive fibrosis.[11–13] In rare cases, the immunological reaction may potentially be the cause of breast implant-associated anaplastic large cell lymphoma (BIA-ALCL); however, our knowledge of these immunological processes is still very limited.[14, 15]

Several studies have investigated the histological characteristics of breast implant capsules, but they are limited by descriptive histological approaches in small, heterogeneous patient populations.[16] The lack of a standardized and reproducible semiquantitative assessment tool for analyzing breast implant capsules prevents an effective comparison of results between studies and thus impairs our ability to build on the knowledge of previous studies. An objective and validated tool for systematically characterizing the histological appearance of the breast implant capsule is needed to create a common language between studies. Such a tool may help us understand why some but not all women develop capsular contracture, and it may be of use in developing new preventive strategies. In this study, we aimed to develop and validate a semiquantitative histological assessment tool that covers the histological characteristics that are seen across a large sample of hematoxylin and eosin (H&E)-stained breast implant capsules. Furthermore, we aimed to show the applicability of the assessment tool by analyzing differences between patients with and without capsular contracture.

## Materials and methods

### Patients and sample material

The histological samples were obtained from a biobank of tissue biopsies excised from the breast implant capsule from patients undergoing exchange or removal of their breast implants after breast augmentation or implant-based breast reconstruction. Patients were included from October 2019 to August 2022 from two plastic surgery departments (*anonymized for peer review*) and three private hospitals (*anonymized for peer review*). The biobank consisted of 400 patients who had their implants removed after breast augmentation and 80 patients who had their implants removed after an implant-based breast reconstruction. Patient and implant characteristics are described in Table 1. Biopsies of approximately 1 cm^2^ were excised from the anterior-inferior part of the fibrous capsule adjacent to the inframammary fold, fixated in a 4% formaldehyde solution and embedded in paraffine. Sections of 3-4 µm were cut and stained with hematoxylin and eosin (H&E). All sections were digitalized using a whole slide scanner and analyzed using NDP.viewer2 (Hamamatsu Photonics).

**Table 1.**
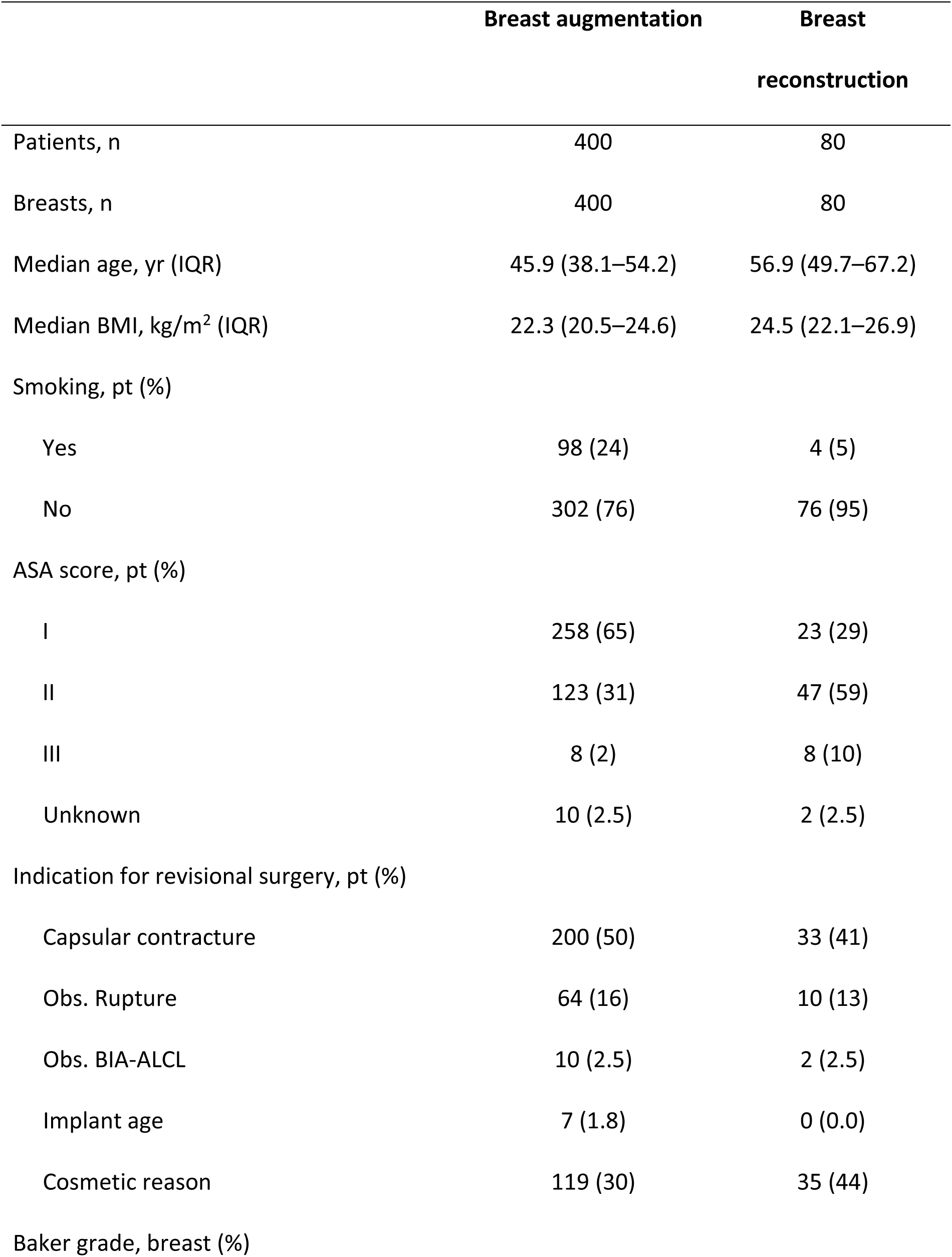
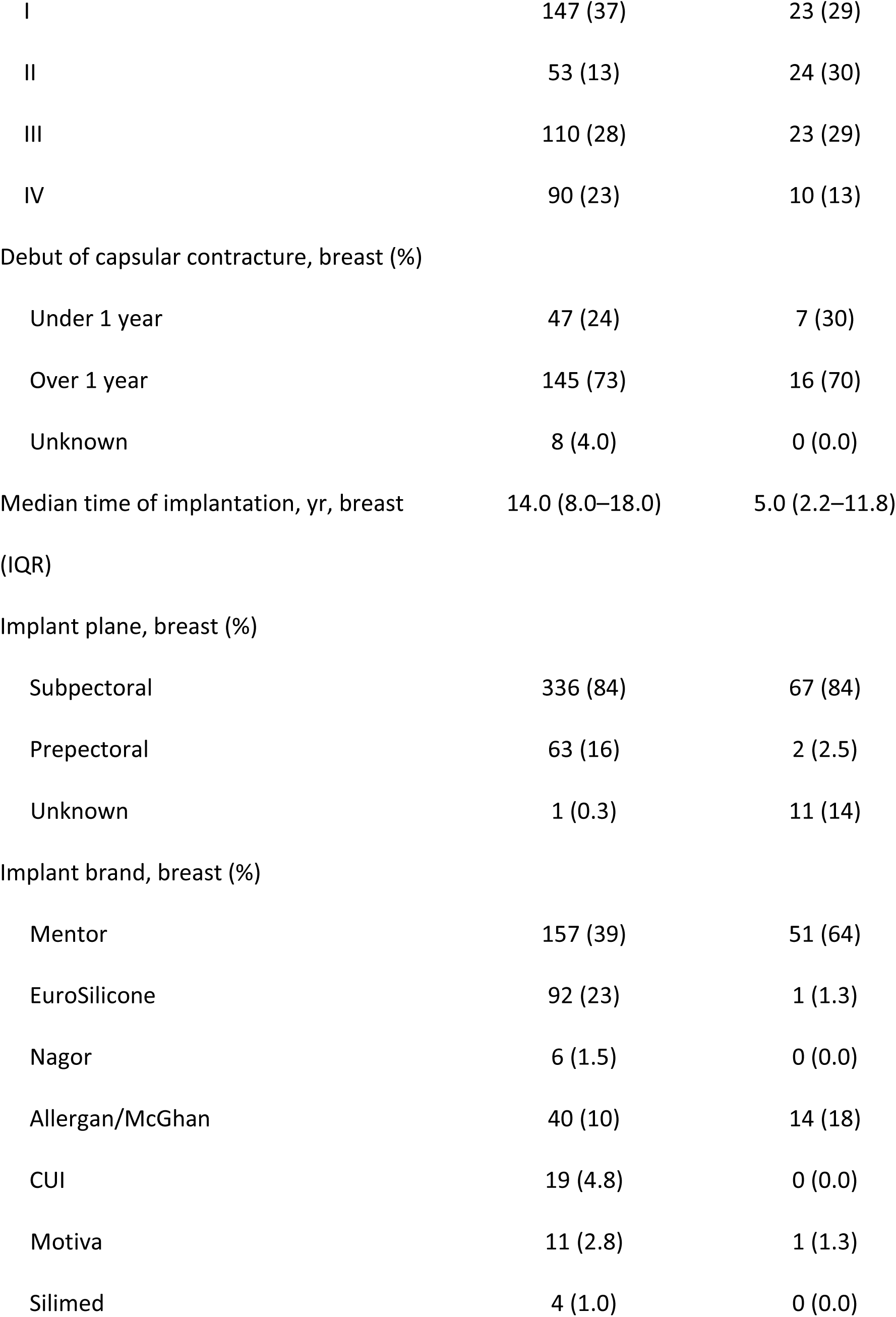
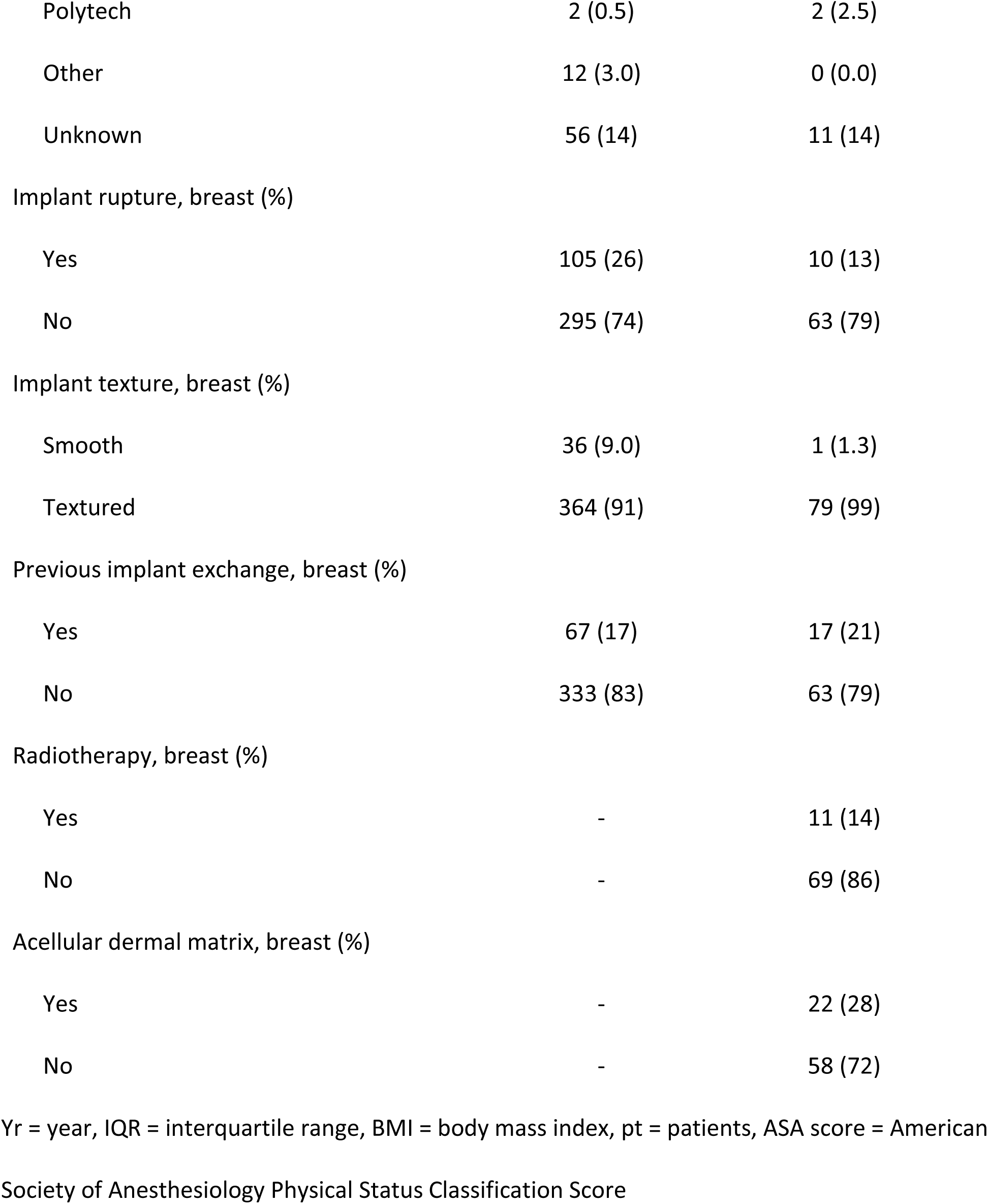
Patient- and implant characteristics.

### Design of the semiquantitative assessment tool

The semiquantitative assessment tool was designed and validated according to principles for valid histologic scoring in research.[17] Potential parameters for the histological assessment tool were chosen by the authors based on a systematic review of histological studies of breast implant capsules.[ref removed for anonymization for peer review] Each selected parameter was transferred to a binary scale or an ordinal scale from absent to severe to assess the prevalence and abundance of each parameter in the biopsies. The reproducibility and applicability of each potential parameter were tested by two observers on breast implant biopsies from 100 randomly selected women from the biobank. Disagreement between observers was discussed with a breast pathologist until a consensus was reached. Parameters with low applicability or rare incidence in the samples were discarded. The ordinal scales were modified to ensure that each scale point was used in the sample. To avoid diagnostic drift, a reference library of histological images illustrating each parameter was developed. The reference library includes detailed instructions on how to assign a semiquantitative score for each parameter. It can be found in Electronic Supplementary Material 1. In the next phase, the assessment tool was validated in terms of observer reproducibility in the samples from the remaining 380 women in the biobank. Two observers scored all 380 samples. In addition, 30 of these samples were scored twice to calculate intraobserver agreement. The biopsies from 100 randomly selected patients were scored by the breast pathologist to assess the agreement between the pathologist and the observers. The results from the histological scorings were used to assess the association between each parameter and the patient’s clinical severity of capsular contracture using the Baker grade. A flowchart of the study design is presented in Figure 1.

**Figure 1.**
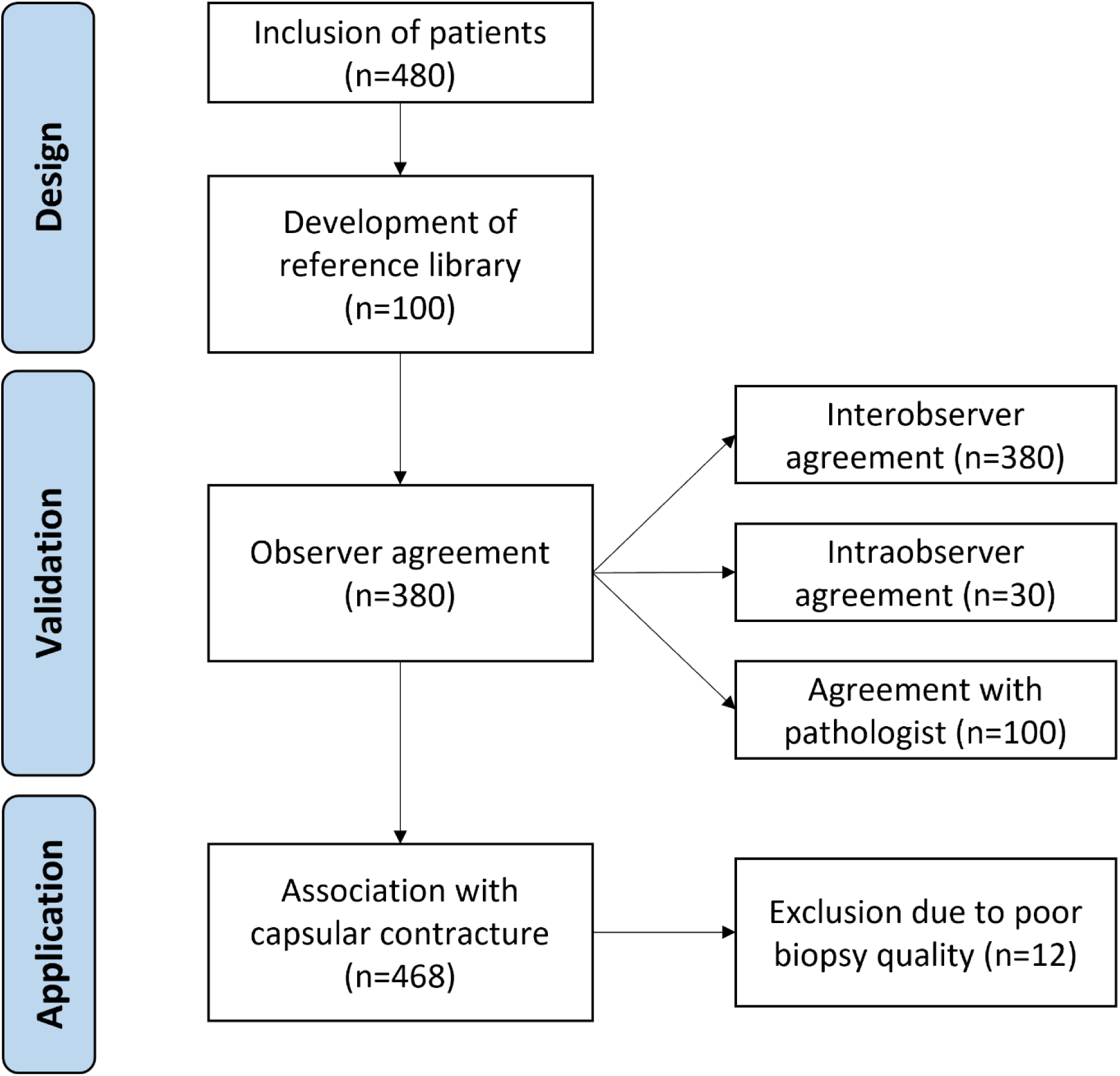
Flow diagram of the study design

### Ethical considerations

Oral and written informed consent was obtained from all patients prior to inclusion in the study in compliance with the Declaration of Helsinki. The study was approved by the Regional Ethics Committee (H-20015276) and the Danish Data Protection Agency (P-2020-176 and P-2020-177).

### Statistics

Agreement between and within observers was calculated with linear weighted Kappa scores. Interobserver agreement compared the scores of each histological parameter between two observers, and intraobserver agreement compared the duplicated scores of each histological parameter. Kappa scores were evaluated as no (0.01-0.20), fair (0.21-0.40), moderate (0.41-0.60), substantial (0.61-0.80) and almost perfect (0.81-1.00) agreement.[18] The association between the Baker grade (Baker I/II vs. Baker III/IV) and the histological parameter was estimated with a multivariable logistic regression with ridge penalization and robust covariance estimation to adjust for clustering of observers. All associations are reported as odds ratios with 95% CIs, and p values were evaluated at an alpha level of 5%. All statistics and plots were performed in R, version 4.2.0.

## Results

### Selection of parameters

We identified 16 potential parameters for the semiquantitative assessment tool. Four parameters were related to fibrosis, including fibroblast and myofibroblast quantification, thickness of the dense collagen layer and organization of the collagen fibers. Four parameters were related to inflammation, including acute inflammation (neutrophil granulocytes), chronic inflammation (lymphocytes), vascularization and calcification. Five potential parameters were related to a foreign-body reaction, including synovial-like metaplasia, free silicone vacuoles, intracellular located silicone in macrophages (foam cells), multinucleated giant cells and granulomatous inflammation (silicone granulomas). Furthermore, the total number of layers in the capsule, activity in the stromal layer and neuromas were identified as potential parameters for the assessment tool.

Based on the initial scoring of the 100 randomly selected biopsies, five parameters were excluded, including acute inflammation, granulomatous inflammation, number of layers, neuromas and histomorphometric grading of synovial-like metaplasia[19] due to low prevalence, low applicability and difficulty to standardize. Ten parameters were included in the final assessment tool, each with its own two-, three- or four-point scale. An overview of the parameters in the assessment tool can be seen in Table 2. The thickness of the collagen layer was divided into four intervals based on the median and interquartile range (IQR) to enable assignment of a semiquantitative score. Reliable morphological distinction between fibroblasts, myofibroblasts and resident macrophages is challenging in H&E-stained sections. Consequently, we chose to combine these into one parameter that evaluates the number of resident cells in the collagen layer. Furthermore, we combined free silicone vacuoles and foam cells into one parameter for silicone infiltration.

**Table 2.**
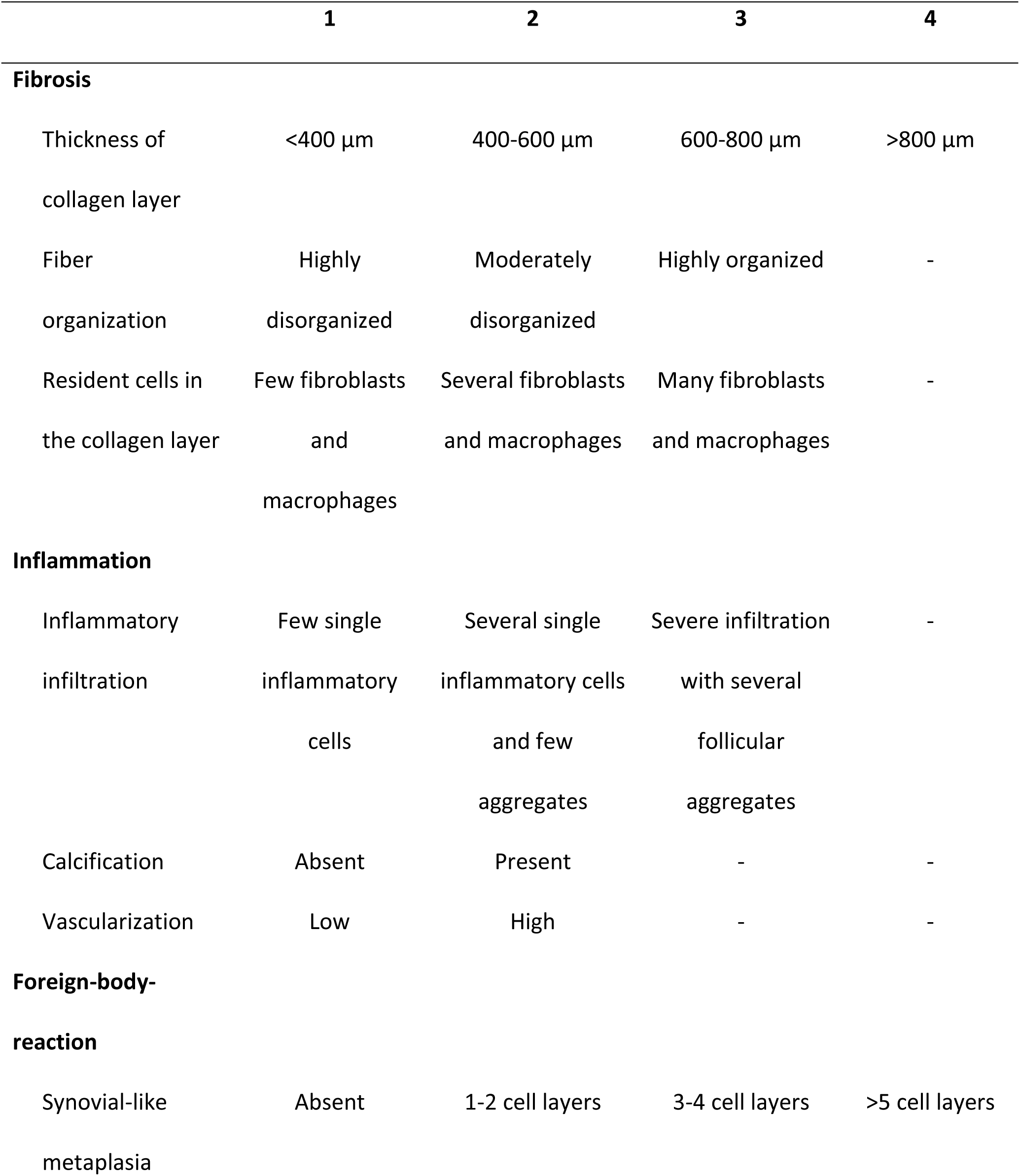
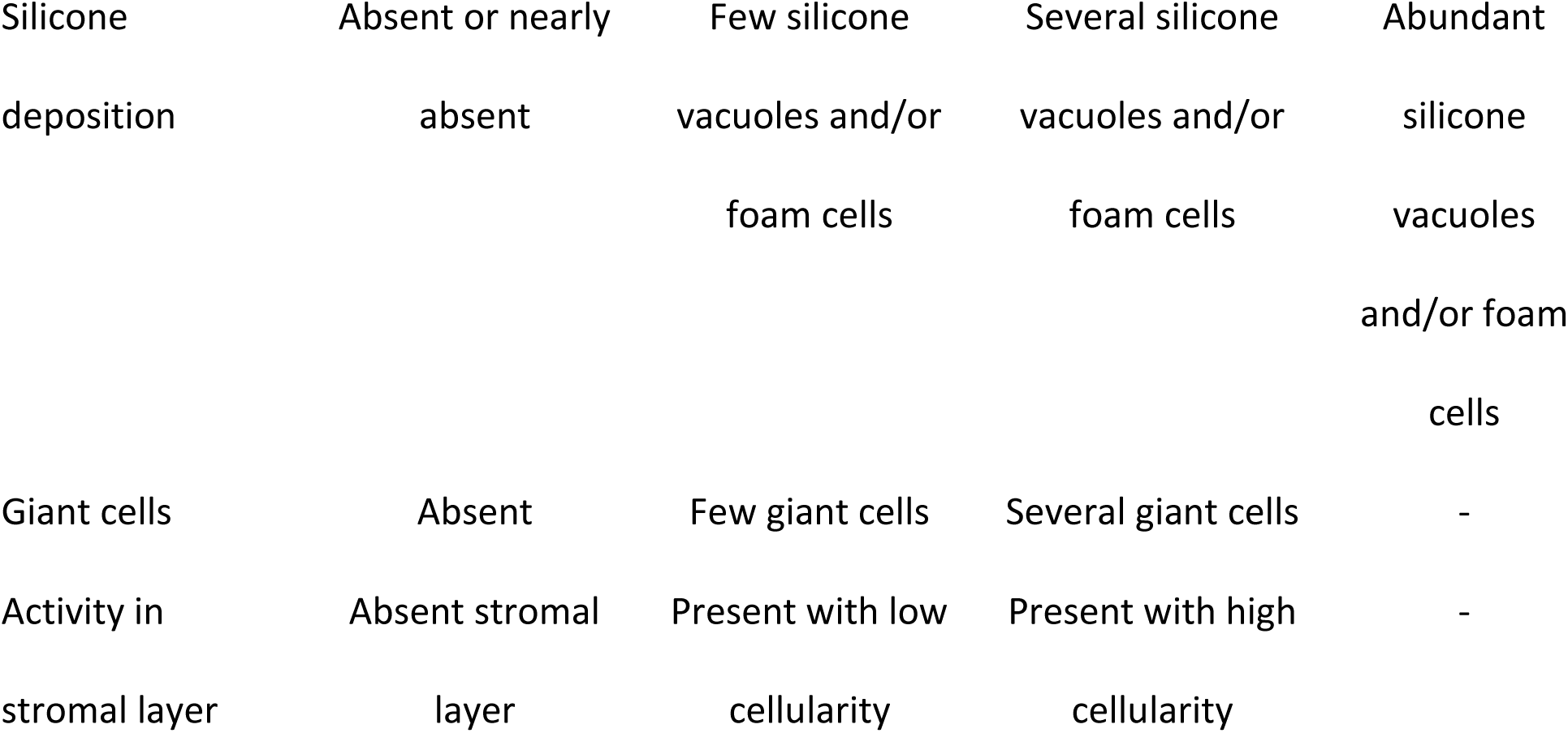
The histological semiquantitative assessment tool for breast implant capsules.

### Reliability and reproducibility of the histological assessment tool

The intra- and interobserver agreements were 0.85 (95% CI 0.85-0.85) and 0.80 (95% CI 0.80-0.80), respectively, which indicate almost perfect agreement. Six biopsies (1.5%) from patients with breast augmentation and six biopsies (7.5%) from patients with breast reconstruction were excluded due to poor quality of the histological slides (for instance, insufficient representation of the full capsule, tissue folding or sectioning artifacts). Examples of poor-quality biopsies have been included in the reference library (ESM1). The agreement between the observers and the breast pathologist was 0.67 (95% CI 0.67-0.67), indicating substantial agreement.

### Histological features of the breast implant capsule

When investigating the fibrotic changes, we found that the collagen layer generally constituted the majority of the implant capsule biopsies and varied greatly in thickness and fiber organization. The thickness of the collagen layer ranged from 72 µm to over 3000 µm. We found that 19% of the samples were highly organized with fibers running parallel to the implant surface and fibroblast nuclei elongated in the direction of traction and parallel to the fibers (Figure 2a). In contrast, 14% of the samples were highly disorganized with multidirectional bundles of fibers and crosscut, rounder fibroblast nuclei. The cellularity of the resident cells in the collagen layer also varied greatly, with 24% of the samples containing many fibrocytes, fibroblasts, myofibroblasts and macrophages, whereas 12% of the samples had a low cellularity with an almost acellular appearance (Figure 2a/2b).

**Figure 2.**
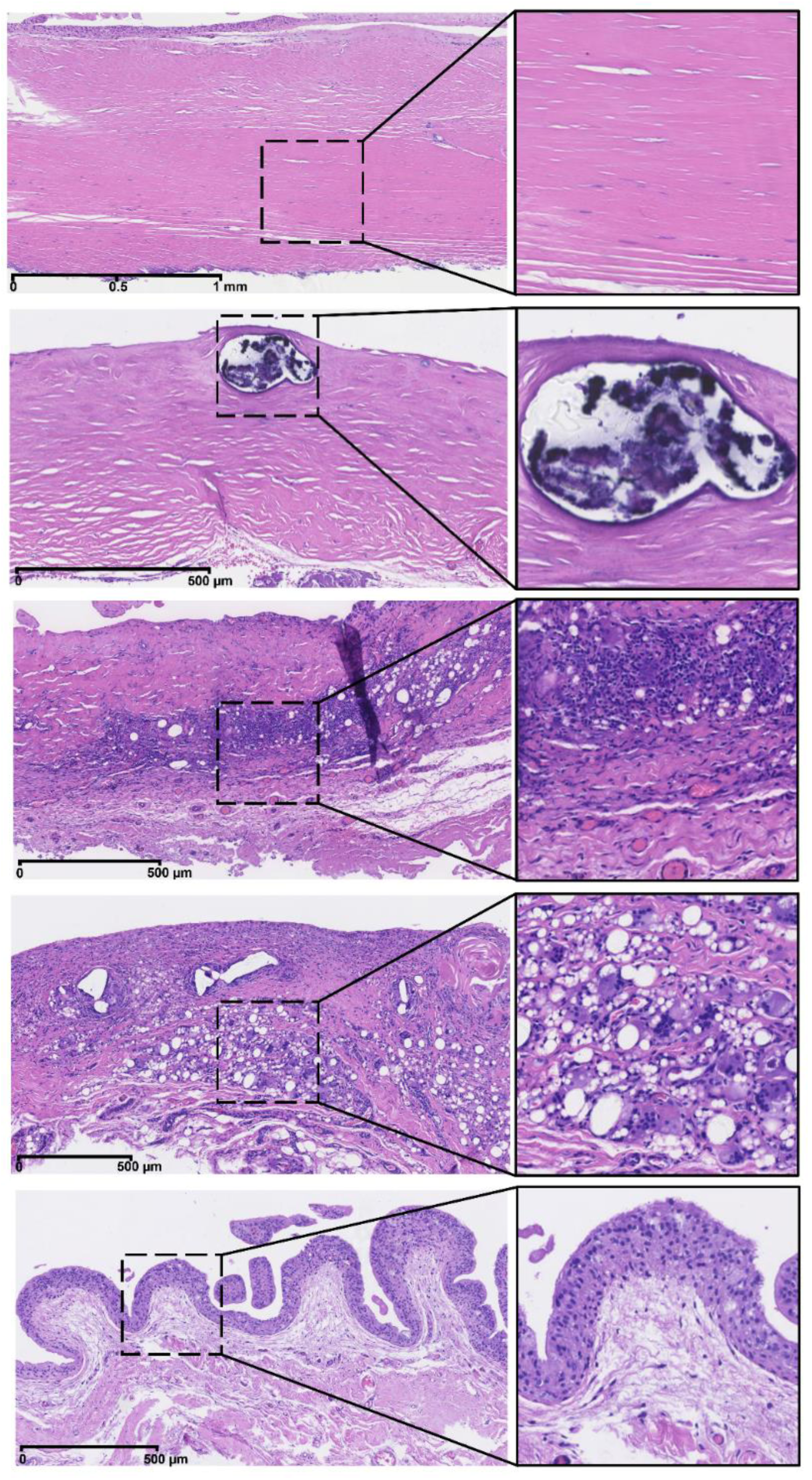
Examples of the histological appearance of breast implant capsules. All scale bars are 500 µm *(a)* capsule with highly organized fibers (3 points) running parallel to the implant surface and fibroblast nuclei elongated in the direction of traction, *(b)* capsule with a low cellularity of resident cells (1 points) and the presence of calcification (2 points), *(c)* capsule with severe inflammatory infiltration (3 points) and high vascularization (2 points), *(d)* capsule with abundant silicone vacuoles and foam cells (4 points) and several multinucleated giant cells (3 points), and *(e)* capsule with synovial-like metaplasia of 3–4 cell layers (3 points) and the presence of a stromal layer with low cellularity (2 points).

When investigating the inflammatory response, we found chronic inflammatory infiltration to be present in 12% of the samples (Figure 2c). Tissue calcification was generally rare and only seen in 6% of the samples. (Figure 2b). Furthermore, we found a low degree of vascularization in most of the biopsies (85%); however, as expected, a high degree of vascularization was often seen in biopsies with severe inflammatory infiltration (Figure 2c).

Silicone was observed extracellularly as small round to irregular translucent droplets of amorphous refractile material as well as intracellularly when phagocytized by histocytes, giving them a foam cell appearance (Figure 2d). Silicone was identified in over half of the included samples (54%). Multinucleated giant cells, which are known to be part of a foreign body reaction, were seen in 22% of the samples and were closely related to the presence of silicone vacuoles and foam cells (Figure 2d). A layer of synovial-like metaplasia was found adjacent to the implant surface in 70% of the samples with a varying thickness from 1 to >5 cell layers (Figure 2e) but was absent in 30% of the samples (Figure 2b). Moreover, we found a layer of stromal tissue to be present between the synovial-like metaplasia and the layer of dense collagen fibers or between bundles of densely packed collagen fibers in 42% of the samples (Figure 2e). See Table 3 for the distributions of histological scores between biopsies from patients with breast augmentation and breast reconstruction.

**Table 3.** Distribution of histological scores between biopsies from patients with breast augmentation and breast reconstruction.

|  | Breast augmentation | Breast reconstruction | Total |
| --- | --- | --- | --- |
| Patients, n | 394 | 74 | 468 |
| **Thickness of collagen layer (%)** |  |  |  |
| <400 µm | 170 (43) | 35 (47) | 205 (44) |
| 400–600 µm | 87 (22) | 14 (19) | 101 (22) |
| 600–800 µm | 51 (13) | 12 (16) | 63 (14) |
| >800 µm | 86 (22) | 13 (18) | 99 (21) |
| **Fiber organization (%)** |  |  |  |
| Highly disorganized | 57 (15) | 7 (9.5) | 64 (14) |
| Moderately disorganized | 255 (65) | 59 (80) | 314 (67) |
| Highly organized | 82 (21) | 8 (11) | 90 (19) |
| **Resident cells in the collagen layer (%)** |  |  |  |
| Few fibroblasts, myofibroblasts and macrophages | 51 (13) | 6 (8.1) | 57 (12) |
| Several fibroblasts and macrophages | 244 (62) | 56 (76) | 300 (64) |
| Many fibroblasts and macrophages | 99 (25) | 12 (16) | 111 (24) |
| **Inflammatory infiltration (%)** |  |  |  |
| Few single inflammatory cells | 348 (88) | 65 (88) | 413 (88) |
| Several single inflammatory cells and few aggregates | 33 (8.4) | 8 (11) | 41 (8.8) |
| Severe infiltration with several follicular aggregates | 13 (3.3) | 1 (1.4) | 14 (3.0) |
| **Calcification (%)** |  |  |  |
| Absent | 365 (93) | 74 (100) | 439 (94) |
| Present | 29 (7.4) | 0 (0.0) | 29 (6.2) |
| **Vascularization (%)** |  |  |  |
| Low | 334 (85) | 63 (85) | 397 (85) |
| High | 60 (15) | 11 (15) | 71 (15) |
| **Synovial-like metaplasia (%)** |  |  |  |
| Absent | 127 (32) | 12 (16) | 139 (30) |
| 1–2 cell layers | 129 (33) | 31 (42) | 160 (34) |
| 3–4 cell layers | 97 (25) | 26 (35) | 123 (26) |
| >5 cell layers | 41 (10) | 5 (6.8) | 46 (9.8) |
| **Silicone deposition (%)** |  |  |  |
| Absent or nearly absent | 172 (44) | 41 (55) | 213 (46) |
| Few silicone vacuoles and/or<br>foam cells | 105 (27) | 21 (28) | 126 (27) |
| Several silicone vacuoles and/or<br>foam cells | 55 (14) | 8 (11) | 63 (14) |
| Abundant silicone vacuoles<br>and/or foam cells | 62 (16) | 4 (5.4) | 66 (14) |
| **Giant cells (%)** |  |  |  |
| Absent | 305 (77) | 61 (82) | 366 (78) |
| Few giant cells | 35 (8.9) | 5 (6.8) | 40 (8.5) |
| Several giant cells | 54 (14) | 8 (11) | 62 (13) |
| **Activity in stromal layer (%)** |  |  |  |
| Absent stromal layer | 238 (60) | 42 (57) | 280 (60) |
| Present with low cellularity | 42 (11) | 7 (9.5) | 49 (11) |
| Present with high cellularity | 114 (29) | 25 (34) | 139 (30) |

### Associations between the histological parameters and capsular contracture

To demonstrate the applicability of the semiquantitative assessment tool, the association between the histological scores and Baker grade was analyzed in 468 patients. We found that four fibrotic and inflammatory parameters were significantly associated with capsular contracture. The thickness of the collagen layer had the strongest association with capsular contracture: 40% of the capsules from breasts with capsular contracture (Baker III/IV) had the highest score with a thickness above 600 µm compared with only 13% of the capsules from breasts without capsular contracture (Baker I/II) (p<0.0001). Moreover, the collagen fibers were significantly more organized with fibers running parallel to the implant surface in capsules from breasts with capsular contracture than in capsules from breasts without capsular contracture (28% vs. 14%, p<0.01).

Infiltration of lymphocytes in the fibrous capsule was also significantly more present in capsules from breasts with capsular contracture than in capsules from breasts without capsular contracture (17% vs. 6%, p<0.01). Furthermore, the presence of calcification was also significantly associated with capsular contracture (14% vs. 0.7%, p<0.001). The remaining parameters were not significantly associated with capsular contracture (Figure 3).

**Figure 3.**
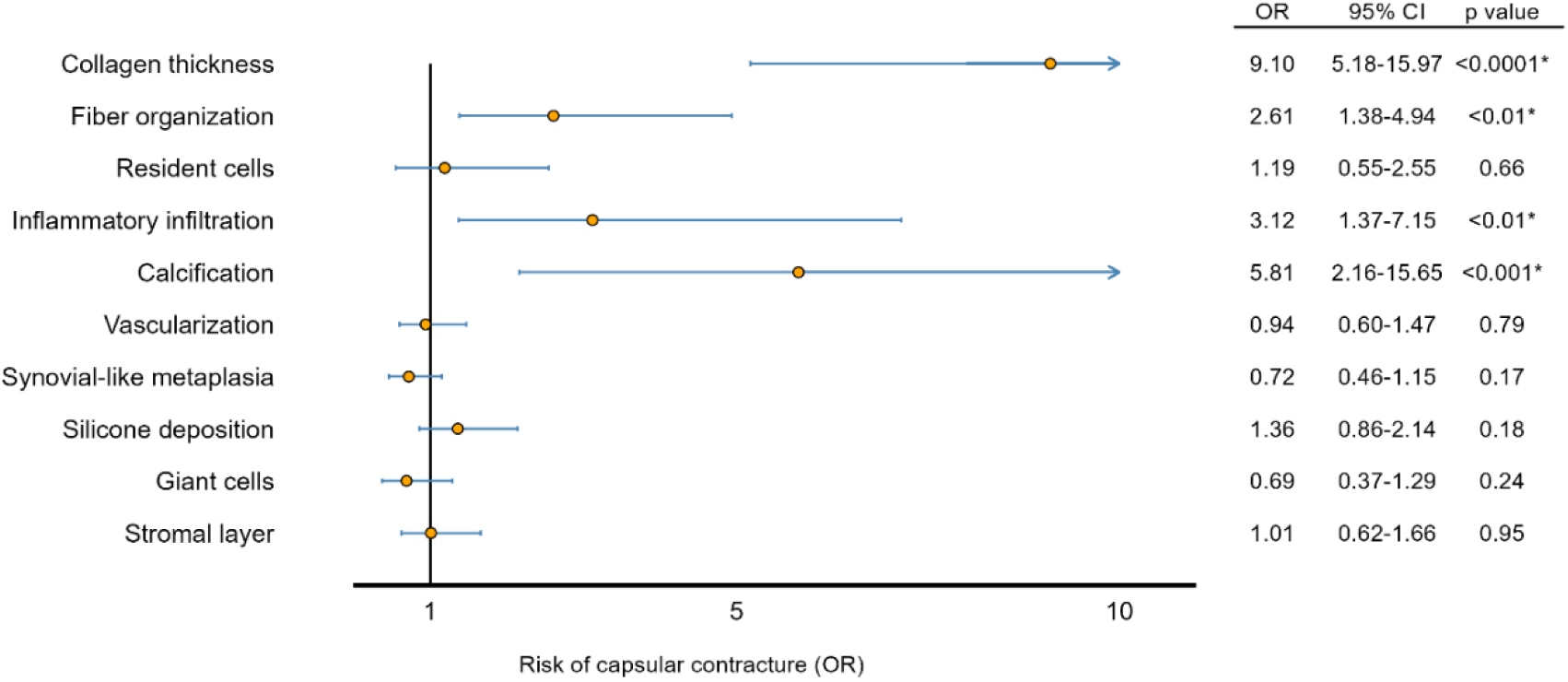
The association between each histological parameter and the clinical degree of capsular contracture estimated with a multivariable logistic regression

## Discussion

To the best of our knowledge, this is the first validated histological semiquantitative assessment tool that can be used to evaluate the immunological and fibrotic response in breast implant capsules. The assessment tool comprises 10 semiquantitative parameters that are highly reliable in terms of intra- and interobserver agreement and with substantial agreement between nonpathologist observers and a breast pathologist. To improve the applicability and usability of the assessment tool, we created a large histological reference index with histological examples of each parameter and thorough instructions on how to score each parameter. We found no acute inflammation or neuromas and only one true silicone granuloma, and consequently, these parameters were discarded from the semiquantitative assessment tool. Our semiquantitative assessment tool provides an objective and standardized measure for characterizing the histology of breast implant capsules. It was validated in 480 patients after breast augmentation or breast reconstruction. The provided distributions of the individual parameters may serve as a strong reference point for future studies. The semiquantitative assessment tool was developed for H&E-stained sections and does not require special immunohistochemical staining, which makes it cost-efficient to include in future studies. The semiquantitative assessment tool can be applied to other important research questions, such as the impact of implant texture, radiotherapy and biological or artificial meshes. Therefore, we recommend that future studies include all 10 parameters and not just those significantly associated with capsular contracture, as we might find other important patterns when including all the visible histological characteristics of breast implant capsules.

As an example of the applicability of the semiquantitative assessment tool, we analyzed the differences between patients with and without capsular contracture and found two fibrotic and two inflammatory parameters significantly associated with capsular contracture. The correlation between capsule thickness and capsular contracture is well established in the literature.[12, 20–23] In a study by Siggelkow et al.[12] analyzing 53 biopsies from 43 patients, a significant association of capsule thickness and Baker grade was found, but they did not find an association with inflammation, silicone deposition or calcification. Bui et al.[21] also found capsules from breasts with capsular contracture to be significantly thicker than those from breasts without capsular contracture based on the analysis of 49 samples from 41 patients. In line with our findings, they found capsules from breasts with capsular contracture to have a significantly higher degree of fiber organization than capsules from breasts without capsular contracture based on vector analysis. Most previous studies investigating the inflammatory reaction to breast implants have either been based on qualitative analysis or have not included statistical tests.[24–26] In a study by Kamel et al.[27] analyzing 93 biopsies, they did, however, find a significant association between the Baker grade and CD3-positive lymphocytes, but they did not find a correlation with calcification. In histological studies of capsular contracture, calcification is described as the end stage of chronic inflammation and the fibrotic reaction.[28, 29] In a study by Peters et al.,[30] calcification was clinically present in 16% of breast implant capsules and was significantly more frequent in first-generation implants[31] than in second- and third-generation implants. In our study, we primarily included biopsies from breasts with textured fourth- or fifth-generation implants and found that calcification was present histologically in 14% of capsules from breasts with capsule contracture and 0.7% of capsules from breasts without capsular contracture. This may be the explanation why previous studies with smaller sample sizes have not been able to find a significant correlation between calcification and capsular contracture.[12, 24, 25, 27] Previous studies have described a loss of cellularity in the fibrous capsule when the time of implantation increases.[29] Myofibroblasts are hypothesized to undergo apoptosis in the later stages of capsular contracture development, similar to what has been observed in wound healing and scar formation.[21] The role of fibroblasts and myofibroblasts in capsular contracture and how the cellularity change over time may be elucidated using specific immunohistochemical stains that improve the distinction between the two cell types, which is not possible in H&E-stained sections.[32–35] Previous studies have found an inverse relationship between the presence of synovial-like metaplasia and capsular contracture and have hypothesized that it plays an important protective role against capsular contracture.[20, 36] Raso et al.[37] hypothesized that the protection is caused by the production of lubricating factors such as proteoglycans, whereas De Bakker[20] et al. hypothesized that synovial-like metaplasia is lost due to the increasing amount of tissue and, thereby, loss of nutrition. Other potential reasons could be that the layer is torn off during implant removal due to a strong adhesion between the implant surface and the capsule, which has been described as a “Velcro effect”[16], or that the layer is lost during sample preparation. The results from this study showed no significant association between the absence of synovial-like metaplasia and capsular contracture. Silicone leakage is believed to trigger capsular contracture by inducing an inflammatory foreign body response.[38, 39] However, we did not find the semiquantitative amount of silicone deposition or the presence of multinucleated giant cells in the capsule to be independently associated with capsular contracture. This could be due to a potential underestimation, as silicone is known to be lost during tissue preparation. Future studies performing a quantitative analysis of silicone leakage adjusted for rupture status may have the sensitivity to find a possible correlation between the amount of silicone in the capsule and capsular contracture.

### Limitations

The study is based on a population of patients among whom the vast majority had textured implants (>90%). This limits the generalizability of the semiquantitative assessment tool in regard to smooth implants. The parameters of the semiquantitative assessment tool are scaled after a population with an even distribution of patients with and without capsular contracture. Therefore, a study that, for example, is concerned with patients with very short implantation times and no capsular contracture will probably find that their patients cluster in one extreme of many of the assessment tool’s parameters. Standardization of the sample site has improved the comparability between samples; however, this might impose a sampling bias, as potential variability of the histological parameters between different locations of the capsule has not been investigated.

## Conclusion

To our knowledge, this is the largest histological study of the breast implant capsule and the morphological changes from capsular contracture. We have developed and validated a histological semiquantitative assessment tool that is highly reliable in terms of agreement and can be used with a standard H&E stain. Thus, it can provide an accurate and objective assessment of breast implant capsules for future research studies and clinical practice. We strongly encourage others to use and perhaps refine this semiquantitative assessment tool.

## Funding information

The study was funded by the Research Fund of Rigshospitalet and the Novo Nordisk Foundation (grant number 052198)

## Conflict of Interest

The authors declare that they have no conflict of interest.

## Ethical approval

All procedures performed in studies involving human participants were in accordance with the ethical standards of the institutional and national research committee and with the 1964 Helsinki declaration and its later amendments or comparable ethical standards.

## Data Availability

All data produced in the present study are available upon reasonable request to the authors

## Electronic Supplementary Material Legend

Electronic Supplementary Material 1. A reference library of histological images illustrating each parameter of the semiquantitative assessment tool with detailed instructions on how to score each parameter.

## Notes

### Competing Interest Statement

The authors have declared no competing interest.

### Funding Statement

This study was funded by the Research Fund of Rigshospitalet and the Novo Nordisk Foundation (grant number 052198)

### Author Declarations

The study was approved by the Regional Ethics Committee (H-20015276) and the Danish Data Protection Agency (P-2020-176 and P-2020-177).

### Summary of Updates

Clarification of title as a histological assessment tool

